# Detection-Guided Artifact Removal for Clinical EEG: A Deep Learning Framework

**DOI:** 10.64898/2026.02.12.26346128

**Authors:** Evans Nyanney, Parthasarathy D. Thirumala, Shyam Visweswaran, Zhaohui Geng

## Abstract

**Objective:** We developed and validated a detection-guided artifact removal framework for clinical electroencephalography (EEG). It corrects only the contaminated segments and preserves artifact-free data.

**Approach:** The framework employs convolutional neural network (CNN) detectors trained on the Temple University Hospital (TUH) Artifact Corpus of 150 recordings from 105 patients. For eye movement artifacts (20 second windows), it uses independent component analysis (ICA) and canonical correlation analysis (CCA). For muscle artifacts (5 second windows), it employs wavelet thresholding and empirical mode decomposition (EMD). For non-physiological artifacts (1 second windows), it utilizes spherical spline interpolation and artifact subspace reconstruction (ASR). Removal is applied exclusively to detector-flagged windows, and unflagged windows remain unchanged. In a held-out test set of 21 patients and 30 recordings, we compared selective and global removal using correlation, root mean squared error (RMSE), and peak signal-to-noise ratio (PSNR).

**Main results:** Selective removal outperformed global removal in all six methods and 18 metric comparisons, with a *p*-value of less than 10^−105^. Selective processing maintained a clean-segment correlation above 0.987, whereas global removal reduced the correlation to values as low as 0.39 for CCA and 0.47 for ASR. CCA removed 74.6% of the eye movement artifact amplitude, EMD removed 99.8% of the high-frequency (30–40 Hz) muscle contamination, and ASR reduced the non-physiological artifact amplitude by 37.1%. The preservation of artifact-free windows remained high for all methods and indicated minimal distortion of the clean EEG.

**Significance:** Detection-guided selective removal addresses a significant limitation of global correction pipelines that can remove clean EEG signals. This framework automates artifact removal without manual review and preserves the signal fidelity for clinical interpretation. Its modular design facilitates integration into real-time monitoring systems for acute and perioperative care.

## 1 Introduction

Continuous scalp electroencephalography (EEG) is central to the detection of seizures, status epilepticus, and cerebral ischemia in acute and perioperative care[1, 2, 3]. Many clinically important events are nonconvulsive and are visible only on the EEG record[4, 5]. Narrative reviews of critical care EEG have reported that real-time monitoring improves seizure detection and supports therapeutic decision-making[6, 7, 8]. Early studies have also shown that continuous EEG reveals electrographic seizures in a substantial fraction of comatose patients[9, 10, 11].

The diagnostic precision and reviewer workload are compromised by physiological artifacts and non-physiological events. Physiological artifacts include eye movements, electromyographic (EMG) activity, chewing, and shivering. Non-physiological events include electrode pops, static, and lead failures[12, 13, 14, 15]. These sources can obscure or mimic pathologies and often require extensive manual cleanup. The foundational work has characterized the spectral and topographic signatures of muscle contamination and other artifact sources[16, 17, 18]. This work motivated automated rejection frameworks such as FASTER[19] and widely adopted ICA-based pipelines[20, 21].

Subsequent developments explored empirical mode decomposition for blink removal[22, 23] and wavelet-based approaches for muscle artifact suppression[24, 25, 26]. Component-based automatic methods include Artifact Subspace Reconstruction[27, 28]. Spatial interpolation methods, such as spherical spline interpolation, have been used to reconstruct signals from corrupted channels[29, 30]. Hybrid multi-method approaches have demonstrated improved performance on heavily contaminated recordings[31, 32]. Many classical methods were trained on small, controlled datasets and showed limited robustness in diverse clinical settings, equipment, and patient populations[12, 13, 33]. Recent deep-learning approaches promise improved performance[34, 35, 36, 37]. Systematic reviews have reported that many algorithms assume homogeneous artifact distributions across channels, subjects, and tasks. This assumption mismatches the heterogeneity of real-world EEG [38, 39, 40, 41].

Many artifact removal pipelines apply corrections globally across entire recordings or apply uniform processing across artifact types[12, 13, 33]. This practice can lead to over-aggressive processing of clean segments or inadequate suppression of contamination[14, 15]. In clinical recordings, artifact families occur on a range of time scales and frequency bands[16, 18]. Transient electrode pops, sustained electromyographic activity[24, 21], and low-frequency ocular events[22, 23] exhibit distinct temporal characteristics. These characteristics suggest that artifact-specific temporal windows and tailored removal strategies can improve removal performance[12, 13, 41]. Component-based methods, such as ASR, can identify nonstationary high-amplitude artifacts[27, 28]. These methods often use uniform thresholds for artifact types[20, 19]. Uniform thresholds can distort clean segments or leave residual contamination[25, 26]. To address this challenge, we previously developed and validated separate lightweight convolutional neural network (CNN) detectors for eye movements, muscle and electromyogram (EMG) artifacts, and non-physiological electrode disturbances. We used the TUH Artifact Corpus with strict patient-level splits[42]. These detectors achieved a specificity of 96% for ocular events (20 s windows), 96% for muscle artifacts (5 s windows) and 98% for electrode artifacts (1 s windows). The F1-scores ranged from 77% to 91% between artifact types. This performance established a validated detection framework that can gate artifact-specific removal algorithms[42].

Based on this validated detection framework[42], we implemented and evaluated artifact removal modules that used detection output to apply targeted correction only to contaminated segments. The modules preserve artifact-free data unchanged[12, 13, 41]. We coupled artifact-specific CNN detectors with previously established removal techniques. We used independent component analysis for ocular artifacts[20, 21, 37], wavelet thresholding for muscle artifacts[43, 24, 25, 26], and spatial interpolation methods for electrode disturbances[29, 30]. Unlike hybrid approaches that apply multiple methods uniformly to all segments[31, 32], our framework activates a removal algorithm only when the corresponding artifact is detected in a class-specific temporal window[42]. This selective activation reduces the processing of clean segments and reduces the risk of signal distortion[14, 33]. We evaluated the effectiveness of removal using two complementary assessments. The assessments were artifact reduction in contaminated segments and signal preservation in clean segments[34, 36]. To ensure clinical safety[15, 9], we conducted preservation studies that showed removal maintained the waveform morphology and spectral content within 2% of the baseline. The studies also showed the preservation of clinically relevant features, such as spike-wave patterns and background rhythms. Quantitative metrics remained stable in representative recordings[43, 44, 35]. This validation is important for clinical applications, as false corrections can mask genuine pathologies[6, 7, 14].

Our objectives are as follows: (1) to implement artifact-specific removal strategies (ICA and CCA for eye movements[20, 45, 37], Wavelet thresholding and EMD for muscle artifacts[43, 24, 25], and interpolation and ASR for electrode disturbances[29, 27, 28]) guided by our previously validated CNN detection models[42]; (2) to evaluate whether detection-guided removal outperforms global removal methods in artifact reduction and signal preservation[12, 13, 41, 33]; (3) to quantify clinical safety through preservation studies on artifact-free segments[43, 44, 15, 14]; and (4) to provide open-source tools and evaluation frameworks for clinical validation of the complete detection-guided removal system[34, 36, 35]. We hypothesized that detection-guided removal would achieve superior artifact reduction and improved preservation of neural signals compared to global removal approaches[38, 39, 40]. Upon completion, we will release the code for the detection-guided removal framework under an open-source license to support future EEG studies and real-time monitoring.

## 2 Methods

### 2.1 Dataset and Participants

For this study, we used the Temple University Hospital (TUH) EEG Artifact Corpus. This corpus is a publicly available dataset of clinical EEG recordings with expert-annotated artifact labels[46, 47]. The corpus contains recordings from patients who underwent routine clinical EEG monitoring between 2002 and 2016 at Temple University Hospital in Philadelphia, Pennsylvania, USA. Recordings were collected across multiple clinical settings. These settings included epilepsy monitoring units, neurointensive care units, and general neurology units. The dataset provides expert annotations for removal evaluation.

The analysis included 150 EEG recordings from 105 unique patients. We maintained the same dataset and patient-level split used in our prior detection work[42]. Patient demographics are not publicly available in the de-identified corpus, but the dataset represents a clinically diverse population under standard neurological monitoring. Monitoring indications included epilepsy evaluation, altered mental status, suspected seizures, and routine neurological assessment. Recording durations varied from 20 minutes to several hours and reflect real-world clinical scenarios.

To prevent data leakage, we divided patients at the individual level into training, validation, and test sets. The training set included 63 patients (approximately 90 recordings, 60%), the validation set included 21 patients (approximately 30 recordings, 20%), and the test set included 21 patients (approximately 30 recordings, 20%). Detection models were developed on the training and validation sets in our prior work[42]. The current removal evaluation was conducted on the held-out test set (21 patients, 30 recordings). This test set was not used for detection training, threshold optimization, or removal parameter tuning. This separation ensured unbiased performance assessment.

Expert neurophysiologists provided ground truth artifact labels. Inter-rater agreement was *κ >* 0.8[47]. The corpus includes 158,884 artifact annotations across 19 categories. For this removal study, we grouped annotations into three primary artifact classes: eye movements (38,004 annotations, 23.9%), muscle-related artifacts that combine electromyographic activity, chewing, and shivering (55,072 annotations, 34.7%), and non-physiological artifacts that combine electrode pops, cable movement, and lead failures (32,158 annotations, 20.2%). Clean segments without artifact annotations served as the reference standard for preservation analysis. These grouped classes align with clinically distinct artifact sources and removal strategies.

All participants in the original TUH corpus underwent EEG monitoring as part of standard clinical care. Informed consent was obtained according to Temple University Hospital protocols. This study used secondary analysis of a publicly available, fully de-identified dataset with no patient identifiers or protected health information. Institutional review board (IRB) approval was not required for the current analysis.

### 2.2 Detection-Guided Artifact Removal Framework

Artifact removal in EEG can be applied globally across an entire recording or selectively to contaminated segments. Global approaches risk distortion of clean neural signals when removal methods assume artifact presence that does not hold uniformly[12, 13]. Selective approaches apply correction only where needed and preserve artifact-free segments. Our framework follows the selective approach. CNN-based detectors[42] identify contaminated windows before application of artifact removal methods (Fig. 1).

**Figure 1:**
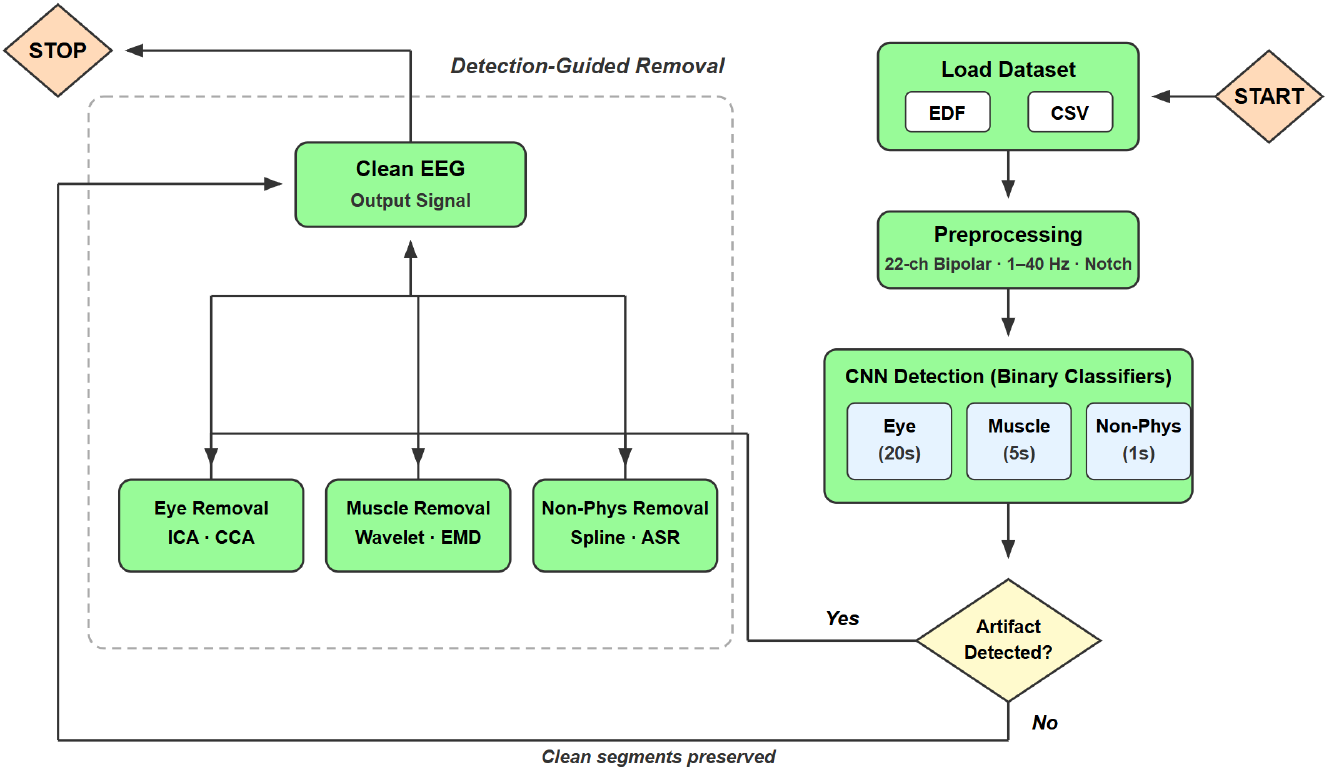
Overview of the detection-guided artifact removal framework. EEG recordings (EDF files) and expert annotations (CSV files) from the TUH Artifact Corpus undergo preprocessing. The preprocessing includes a 22-channel bipolar montage, a 1–40 Hz bandpass filter, 50/60 Hz notch filters, and RobustScaler normalization. Convolutional neural network (CNN) classifiers detect three artifact types: eye movement (20 s windows), muscle (5 s windows), and non-physiological (Non-Phys, 1 s windows). Detected artifacts are processed by type-specific methods. The methods include ICA/CCA for eye movement, Wavelet thresholding/EMD for muscle artifacts, and spherical spline interpolation/ASR for non-physiological artifacts. Clean segments bypass removal and remain unchanged.

For detection, we used binary classifiers trained in our prior work[42] for three artifact types: eye movements, muscle artifacts, and non-physiological artifacts. Each classifier processes an EEG window and outputs a probability. We converted this probability to a binary decision with fixed thresholds (eye: 0.355, muscle: 0.418, non-physiological: 0.359). Thresholds were selected with Youden’s *J* statistic. Detection window lengths were 20 s for eye movements, 5 s for muscle artifacts, and 1 s for non-physiological artifacts[42]. On the test set, detector-flagged segments showed *>*95% agreement with ground truth labels from expert annotators.

For removal, we implemented two established methods for each artifact type to evaluate generalizability. For eye artifacts, we used ICA[20] and CCA[45]. For muscle artifacts, we used Wavelet thresholding[43] and EMD[24]. For non-physiological artifacts, we used spherical spline interpolation[29] and ASR[27]. Each method was applied only to windows flagged by the corresponding detector. Clean windows bypassed all processing and preserved the original signals[12, 13, 33].

### 2.3 EEG Preprocessing

Data preprocessing followed the protocol from our detection study[42]. Original EEG recordings in the TUH corpus used standard clinical equipment with the International 10–20 electrode placement system. Recordings contained 20–36 channels sampled at 250–1000 Hz. We resampled all recordings to 250 Hz with polyphase filtering. This resampling standardized the temporal resolution across recordings.

We converted recordings to a standardized 22-channel bipolar montage in the double-banana configuration. The montage included five chains: left temporal (FP1-F7, F7-T3, T3-T5, T5-O1), right temporal (FP2-F8, F8-T4, T4-T6, T6-O2), central (A1-T3, T3-C3, C3-CZ, CZ-C4, C4-T4, T4-A2), left parasagittal (FP1-F3, F3-C3, C3-P3, P3-O1), and right parasagittal (FP2-F4, F4-C4, C4-P4, P4-O2). This montage provides spatial coverage and supports standard clinical interpretation.

Signals were bandpass filtered between 1 and 40 Hz to remove low-frequency drift and high-frequency noise. Notch filters at 50 and 60 Hz removed power line interference. The 1–40 Hz range emphasizes cerebral activity and reduces contamination from high-frequency muscle artifacts above 50 Hz. After filtering, we re-referenced signals to the common average. We also removed the DC offset by subtracting the channel mean from each channel.

In amplitude normalization, RobustScaler was utilized independently for each channel. RobustScaler standardizes features by removing the median and scaling by the interquartile range. This approach reduces sensitivity to outliers and extreme amplitude values common in clinical EEG. These preprocessing steps produced uniform input data for the detection and removal stages.

### 2.4 Eye Movement Artifact Removal

Eye movement artifacts originate from ocular dipole activity and affect frontal electrodes with characteristic low-frequency signatures below 4 Hz[20, 16, 18]. For eye artifact removal, we implemented two decomposition-based methods: independent component analysis (ICA) and canonical correlation analysis (CCA)[45, 37]. Both methods decompose the EEG signal to isolate ocular activity from neural signals. Windows classified as clean by the CNN detector are not processed and retain their original signals[42].

#### 2.4.1 Independent Component Analysis

Independent component analysis (ICA) decomposes multichannel EEG into statistically independent source components[20, 21]. For ICA, we used FastICA with the logcosh contrast function (max iterations = 800, tolerance = 0.0005, parallel algorithm with unit-variance whitening)[20]. The number of independent components equals the number of channels (22). The model is fitted on artifact windows identified by the CNN detector[42].

For component identification, we used four criteria derived from known properties of eye artifacts[22, 23]: (1) correlation with the detection mask above 0.20, (2) frontal dominance ratio ≥ 0.75 based on mixing-matrix weights at frontal channels relative to all channels[16], (3) kurtosis above 1.0 to indicate a super-Gaussian distribution characteristic of transient eye movements[20, 25], and (4) a low-frequency to high-frequency power ratio above 1.2. The power ratio was computed as spectral power in 0.1–4 Hz relative to 8–30 Hz with Welch’s method[18]. Components that satisfy all four criteria are identified as eye artifacts[37]. Selection is limited to the single highest-correlated component to prevent over-removal[12, 13].

We applied soft attenuation rather than the complete removal of identified components. The base attenuation factor was 0.6 (40% amplitude reduction). This approach preserves residual neural activity that may overlap with ocular sources[21, 30]. The attenuation factor varies with detection confidence. Higher detection probabilities produce greater attenuation, whereas probabilities near the threshold receive more conservative correction. Soft attenuation reduces the risk of signal distortion associated with complete component removal[12]. The cleaned signal is reconstructed from the attenuated components. Artifact removal is applied only to windows flagged by the CNN detector[42]. Clean windows preserve original signals exactly.

#### 2.4.2 Canonical Correlation Analysis

Canonical correlation analysis (CCA) identifies linear combinations of channel groups that maximize mutual correlation[45]. The method uses the frontal distribution of eye artifacts. For CCA, we partitioned the 22 channels into frontal (4 channels: FP1-F7, FP2-F8, FP1-F3, FP2-F4) and non-frontal (18 channels) groups[16]. Four canonical variates (max iterations = 500) extract shared activity between these regions[45].

The canonical variates represent directions of maximum correlation between frontal and non-frontal channel groups. The first variate extracts the most prominent shared signal. This signal typically reflects ocular activity that propagates from frontal to posterior regions[22]. For component identification, we used correlation with the detection mask. Components above a threshold of 0.25 are marked as eye-related[45]. Selection is limited to the top two highest-correlated components to prevent over-removal. Identified components undergo soft attenuation with a base factor of 0.6. Adaptive scaling is applied based on detection confidence[30, 26]. Artifact removal is applied only to windows flagged by the CNN detector[42]. Clean windows remain unmodified.

### 2.5 Muscle Artifact Removal

Muscle artifacts arise from electromyographic (EMG) activity and contaminate EEG in high-frequency bands above 20 Hz[43, 16, 18, 24]. For muscle artifact removal, we implemented two frequency-based methods: wavelet thresholding and empirical mode decomposition (EMD)[25, 23]. Both methods target high-frequency components where EMG activity is concentrated. Artifact removal is applied only to windows flagged by the CNN detector[42]. Windows classified as clean retain their original signals without modification[42].

#### 2.5.1 Wavelet Thresholding

Wavelet denoising exploits the sparse representation of EMG activity in the wavelet domain to separate muscle contamination from neural signals[43, 26, 25]. For wavelet thresholding, we used the Daubechies-4 (db4) wavelet with four-level decomposition[43, 30]. The removal process combines low-pass filtering with adaptive wavelet thresholding. A fourth-order Butterworth low-pass filter with a 30 Hz cutoff attenuates high-frequency EMG components[16, 18].

The filtered signal undergoes wavelet decomposition. Detail coefficients are thresholded with BayesShrink[25, 26]. BayesShrink is an adaptive method that estimates threshold values from the noise variance of each subband. For detail levels 1–3 (highest frequencies), BayesShrink computes the threshold as 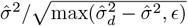, where 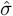 is the median absolute deviation of detail coefficients divided by 0.6745, 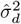 is the variance of the detail coefficients, and *ϵ* is a small constant for numerical stability[43]. For lower detail levels, the universal threshold 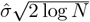 is applied, where *N* is the coefficient length[25].

All thresholding uses soft shrinkage to avoid discontinuities[26]. A safeguard prevents over-attenuation. If the cleaned signal standard deviation falls below 30% of the original, the output is blended with the low-pass filtered signal[12, 13]. The safeguard ensures that excessive signal reduction does not occur. The cleaned signal retains neural activity while EMG contamination is reduced.

#### 2.5.2 Empirical Mode Decomposition

Empirical mode decomposition (EMD) decomposes signals into intrinsic mode functions (IMFs) ordered from highest to lowest frequency[24, 23]. Muscle artifacts are located mainly in the first few IMFs. For EMD, we used a maximum iteration count of 1000 and extracted up to 8 IMFs per channel[24]. The two highest-frequency IMFs are removed, and the signal is reconstructed from the remaining IMFs.

The reconstruction preserves lower-frequency neural activity and attenuates broadband EMG contamination[24, 21]. If EMD produces fewer than two IMFs for a given signal segment, the original signal is retained[12]. The fallback ensures that failed decomposition does not corrupt the output.

### 2.6 Non-Physiological Artifact Removal

Non-physiological artifacts include electrode pops, cable movement, and lead failures that produce transient high-amplitude deflections or sustained signal distortions[12, 13, 14, 15]. Unlike physiological artifacts with predictable spatial and spectral properties[16, 18], non-physiological artifacts can affect any channel and can present with variable morphology[33]. For non-physiological artifact removal, we implemented two spatial reconstruction methods: spherical spline interpolation and artifact subspace reconstruction (ASR)[28]. Artifact removal is applied only to windows flagged by the CNN detector[42], and windows classified as clean retain their original signals without modification[42].

#### 2.6.1 Spherical Spline Interpolation

Spherical spline interpolation reconstructs corrupted channels from the spatial pattern of neighboring good channels[29, 48, 30]. For bad channel identification, we used two complementary criteria. The first criterion computes the Z-score of each channel’s standard deviation relative to the baseline distribution from clean windows, and channels above 2.0 are marked as bad[29]. The second criterion computes the Z-score of maximum amplitude to capture transient spikes, and channels above 1.8 are marked as bad.

Channels flagged by either criterion are interpolated. For interpolation, we computed 3D coordinates for each bipolar channel on a unit sphere. The coordinates were obtained by averaging the 10–20 electrode positions of the constituent electrodes[48]. The interpolation uses a Gaussian kernel approximation to the spherical spline basis functions with *σ* = 0.5 and a regularization parameter *λ* = 10^−3^ for numerical stability[29, 30]. Bad channel values are reconstructed as weighted combinations of good channel values, and the weights are determined by spatial proximity on the scalp surface[48].

If the matrix solution fails, the method falls back to inverse distance weighting[29]. If fewer than three good channels remain, the method falls back to mean replacement[12]. Windows with excessive contamination (more than 50% of channels affected) are rejected, and a clean window from the same recording is substituted[13]. For windows that the detector flags as non-physiological artifacts but for which no specific bad channels are identified, temporal median filtering (kernel size 15) is applied. Outlier removal uses a threshold of 1.8 times the median absolute deviation, and Gaussian smoothing uses *σ* = 4.0[12]. For interpolated channels, median filtering (kernel size 7) and Gaussian smoothing (*σ* = 3.5) are applied. For other channels in interpolated windows, median filtering (kernel size 5) and Gaussian smoothing (*σ* = 2.5) are applied to suppress residual artifacts.

#### 2.6.2 Artifact Subspace Reconstruction

Artifact subspace reconstruction (ASR) identifies and attenuates high-variance signal components that deviate from a learned baseline distribution[27, 44, 28]. From clean windows identified by the detector, we computed a reference covariance matrix. Eigenvalue decomposition of the baseline covariance produces whitening and coloring transformation matrices[44]. The whitening matrix transforms the signal to have unit variance in all directions.

For each artifact window, the signal is centered by subtraction of the baseline mean and then whitened with the baseline covariance[27]. Components with variance above 2.5 standard deviations (burst threshold) or peak amplitude above 2.0 times the reference peak are identified as artifacts[44, 28]. Exceeding components are scaled down so that component variance matches the threshold. More aggressive scaling is applied for larger deviations (down to 0.15 for moderate artifacts and 0.05 for extreme artifacts)[27]. The signal is reconstructed through the inverse coloring transformation[27]. Gaussian smoothing (*σ* = 1.5) is applied after reconstruction to suppress residual artifacts[12, 13]. The method attenuates transient high-amplitude components while preserving the overall signal structure[12, 13].

### 2.7 Statistical Analysis and Performance Evaluation

#### Statistical Analysis

To measure the benefit of detection-guided removal, we compared per-window RMSE on clean data between selective and global removal strategies. For each removal method (independent component analysis (ICA), canonical correlation analysis (CCA), Wavelet thresholding, empirical mode decomposition (EMD), spherical spline interpolation, and artifact subspace reconstruction (ASR)), we computed RMSE for each clean window under both approaches. The paired design treated each window as its own control. This design enables a direct comparison between the two removal strategies.

For normality assessment, we used Shapiro–Wilk tests on per-window RMSE values from clean windows. The tests indicated non-normal distributions for all methods (Shapiro–Wilk *p <* 0.05 for selective RMSE, global RMSE, and paired differences across all six removal methods). These results supported the use of non-parametric tests.

To test whether selective removal produced systematically lower RMSE than global removal, we used the Wilcoxon signed-rank test. The Wilcoxon test is distribution-free and robust to outliers. We specified a one-sided alternative hypothesis (*H*_1_: selective RMSE *<* global RMSE). This test assessed whether detection-guided removal better preserves clean data.

The test statistic equals the minimum of *T* ^+^ (sum of ranks for positive differences) and *T*^−^ (sum of ranks for negative differences). The differences were defined as selective RMSE minus global RMSE. We considered *p*-values *<* 0.05 as statistically significant. For each method, we computed the proportion of windows for which selective removal produced lower RMSE than global removal. This proportion provides a descriptive measure of improvement consistency. Results showed statistically significant improvements for all methods (*p*-values between 2.54 *×* 10^−105^ and 4.42 *×* 10^−116^). These results indicate that detection-guided selective removal better preserves clean EEG segments than global application of removal methods.

#### Performance Evaluation

On the held-out test set (21 patients, 30 recordings), we evaluated removal quality with ground truth artifact labels from expert annotators. Two aspects were assessed: preservation of clean data and reduction of artifact content.

For preservation metrics, we used peak signal-to-noise ratio (PSNR) for all artifact types. PSNR measures preservation quality relative to peak signal amplitude. This metric is appropriate for both continuous contamination and transient spikes. Eye movement and muscle artifacts produce contamination over extended durations, and peak amplitude captures the window-level dynamic range. Non-physiological artifacts produce transient high-amplitude spikes, and peak amplitude captures the magnitude of these events. Preservation relative to peak amplitude quantifies signal fidelity for both artifact types.

In artifact-free windows, we computed root mean squared error (RMSE) to measure the difference between original signal *x* and the cleaned signal 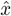 as

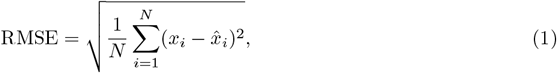

where *N* is the number of samples, *x*_*i*_ is the *i*-th sample of the original signal, and 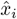 is the corresponding sample after processing. Lower values indicate better preservation of clean segments.

The Pearson correlation coefficient *r* between original and cleaned signals was computed in artifact-free windows. Values near one indicate close agreement between cleaned and original signals and confirm minimal distortion.

For all artifact types, we computed PSNR to quantify preservation quality in decibels as

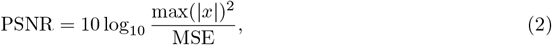

where max(|*x*|) is the maximum absolute amplitude of the original signal and 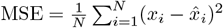 is the mean squared error between original and cleaned signals. Higher values indicate better preservation.

In artifact-contaminated windows, we computed the percentage decrease in signal amplitude as

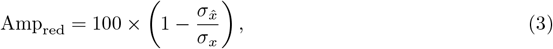

where *σ*_*x*_ and 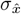 are the standard deviations of the original and cleaned signals in artifact windows. Higher values indicate greater artifact suppression. For eye movement artifacts, we measured amplitude reduction in frontal bipolar channels (FP1-F7, FP2-F8, FP1-F3, FP2-F4), since ocular contamination is most prominent in these channels. For muscle artifacts, we measured spectral power reduction in the 30–40 Hz band, since electromyographic activity predominates in this range. For non-physiological artifacts, we measured peak amplitude reduction alongside overall amplitude reduction. These metrics evaluate artifact removal performance independent of artifact detection performance. Detection performance was validated in our prior work[42].

## 3 Results

### 3.1 Artifact Removal Performance

#### Eye Movement Artifact Removal

To evaluate eye movement artifact removal performance, we tested ICA and CCA on the held-out test set (21 patients, 30 recordings). The CNN detector identified 512 artifact windows from 1303 total windows and achieved an F1-score of 0.895 (precision 0.971, recall 0.831) (Table 1). The precision of 0.971 indicates that most detected windows contain true artifacts, and the recall of 0.831 indicates that the detector identifies most artifact-contaminated segments. The F1-score of 0.895 indicates a balanced trade-off between precision and recall for eye movement detection.

**Table 1:** Eye movement artifact removal summary on held-out test windows.

| Metric | ICA | CCA | $\Delta$<br>(CCA-ICA) |
| --- | --- | --- | --- |
| | $n = 1303$ (598 artifact, 705 clean) | $n = 1303$ (598 artifact, 705 clean) | |
|  | *Detected artifacts: 512* | *Detected artifacts: 512* |  |
|  | *Removed comps: 1 ([20])* | *Removed comps: 2 ([2,3])* |  |
| Detection performance |  |  |  |
| Precision | 0.971 | 0.971 | 0.000 |
| Recall | 0.831 | 0.831 | 0.000 |
| F1-score | 0.895 | 0.895 | 0.000 |
| Clean preservation (clean windows, $n = 705$) | | | |
| PSNR (dB) | **84.15** | 80.44 | -3.71 |
| RMSE ($\mu V$) | **0.0827** | 0.1267 | +0.0440 |
| Correlation | **0.9997** | 0.9992 | -0.0005 |
| Artifact reduction (true artifact windows, $n = 598$) | | | |
| Amplitude reduction (%) | 35.0 | **74.6** | +39.6 |
| Frontal reduction (%) | 45.7 | **67.1** | +21.4 |

**Table description:** Total windows: 1303 (598 ground-truth artifact, 705 clean). Window length = 20 s (5000 samples at 250 Hz). Removal is applied only to detected windows (no global correction). $\Delta$ reports CCA-ICA. For RMSE.

For clean window preservation (*n* = 705), we evaluated both methods. ICA showed correlation of 0.9997, RMSE of 0.0827 *µ*V, and PSNR of 84.15 dB, whereas CCA showed correlation of 0.9992, RMSE of 0.1267 *µ*V, and PSNR of 80.44 dB (Table 1). Both methods showed correlation values above 0.999. These results indicate that the cleaned signals match the original clean segments with minimal distortion. The differences in PSNR (3.71 dB) and RMSE (0.0440 *µ*V) were small, and both approaches preserved clean segments effectively.

For artifact reduction on true artifact windows (*n* = 598), we assessed both methods. ICA achieved 35.0% amplitude reduction and 45.7% frontal channel reduction, whereas CCA achieved 74.6% amplitude reduction and 67.1% frontal channel reduction (Table 1). The 39.6% difference in amplitude reduction (CCA −ICA) reflects CCA’s ability to extract correlated components between frontal and non-frontal channels. CCA achieved 21.4% greater frontal channel reduction than ICA. The frontal channels (FP1-F7, FP2-F8, FP1-F3, FP2-F4) represent the region where ocular contamination is most prominent.

The difference in overall amplitude reduction (74.6% for CCA versus 35.0% for ICA) suggests that maximization of correlation between frontal and non-frontal channels supports more complete artifact removal across the signal. Figures 2 and 3 show representative removal results for both methods.

**Figure 2:**
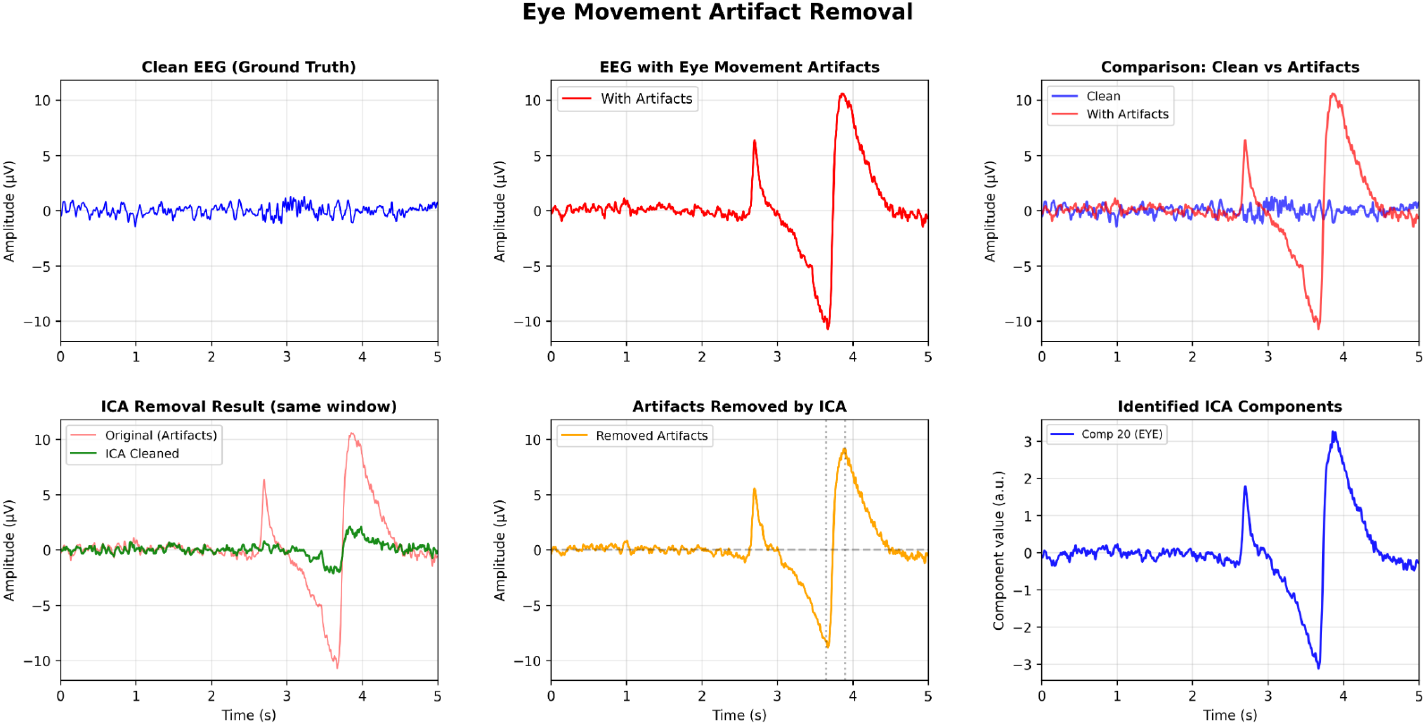
ICA eye movement artifact removal: (top row) clean EEG reference window (separate artifact-free example), artifact-contaminated EEG, and comparison overlay; (bottom row) ICA removal result (original vs cleaned) from the same detected artifact window, removed artifactual components, and identified ICA component 20 (eye artifact).

**Figure 3:**
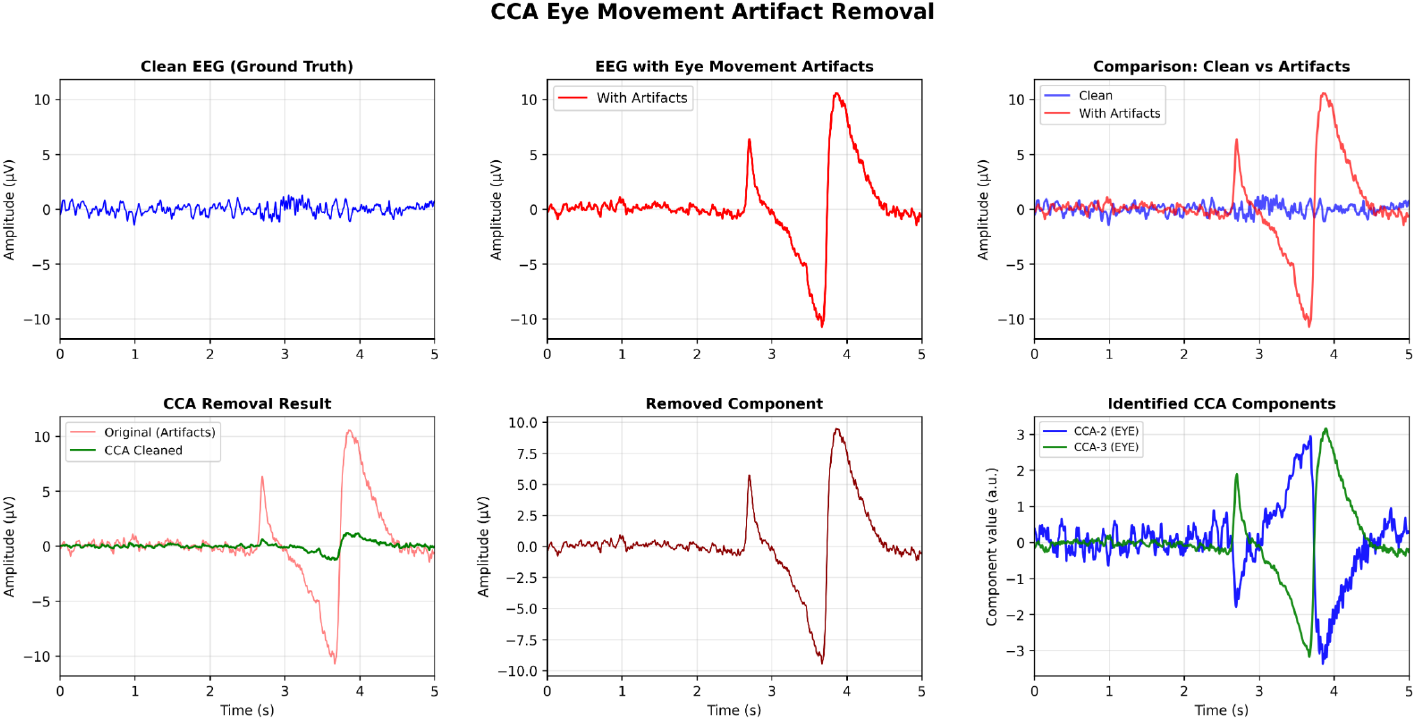
CCA eye movement artifact removal: (top row) clean EEG reference window (separate artifact-free example), artifact-contaminated EEG, and comparison overlay; (bottom row) CCA removal result (original vs cleaned) from the same detected artifact window, removed artifactual components, and identified CCA components 2 and 3 (eye artifacts).

#### Muscle Artifact Removal

To evaluate muscle artifact removal performance, we tested Wavelet thresholding and EMD on the held-out test set (21 patients, 30 recordings). The CNN detector identified 374 artifact windows for Wavelet thresholding and 303 artifact windows for EMD from 1098 total windows. The detector achieved F1-scores of 0.884 (precision 0.906, recall 0.863) for Wavelet thresholding and 0.848 (precision 0.974, recall 0.751) for EMD (Table 2). EMD achieved higher precision than Wavelet thresholding (0.974 versus 0.906). This difference indicates fewer false positive detections for EMD. Wavelet thresholding achieved a higher recall than EMD (0.863 and 0.751). This difference indicates that Wavelet thresholding identified a larger fraction of true artifact windows. The F1-scores indicate a balanced trade-off between precision and recall for both methods.

**Table 2:** Muscle artifact removal summary on held-out test windows.

| Metric | Wavelet | EMD | $\Delta$<br>(EMD– Wavelet) |
| --- | --- | --- | --- |
| | $n = 1098$ (393 artifact, 705 clean) | $n = 1098$ (393 artifact, 705 clean) | |
|  | Detected artifacts: 374 | Detected artifacts: 303 |  |
|  | True artifacts: 393 | True artifacts: 393 |  |
|  |  | IMFs removed: 2 |  |
| Detection performance |  |  |  |
| Precision | 0.906 | **0.974** | +0.068 |
| Recall | **0.863** | 0.751 | -0.112 |
| F1-score | **0.884** | 0.848 | -0.036 |
| Clean preservation (clean windows, $n = 705$) | | | |
| PSNR (dB) | 79.58 | **82.87** | +3.29 |
| RMSE (μV) | 0.1399 | **0.0958** | -0.0441 |
| Correlation | 0.9990 | **0.9995** | +0.0005 |
| Artifact reduction (true artifact windows, $n = 393$) | | | |
| Amplitude reduction (%) | **18.8** | 17.6 | -1.2 |
| HF reduction (30–40 Hz) (%) | 97.9 | **99.8** | +1.9 |
| Beta reduction (20–30 Hz) (%) | 87.0 | **99.1** | +12.1 |

**Table description:** Total windows: 1098 (393 ground-truth artifact, 705 clean). Window length = 5 s (1250 samples at 250 Hz). Removal is applied only to detected windows (no global correction). Wavelet: low-pass filter (30 Hz) + BayesShrink wavelet denoising. EMD: empirical mode decomposition removing top 2 IMFs. $\Delta$ reports EMD – Wavelet. For RMSE.

For clean window preservation (*n* = 705), we evaluated both methods. Wavelet thresholding showed correlation of 0.9990, RMSE of 0.1399 *µ*V, and PSNR of 79.58 dB, whereas EMD showed correlation of 0.9995, RMSE of 0.0958 *µ*V, and PSNR of 82.87 dB (Table 2). Both methods showed correlation values above 0.999. These results indicate that the cleaned signals match the original clean segments with minimal distortion. The differences in PSNR (3.29 dB) and RMSE (0.0441 *µ*V) indicate that EMD preserved clean segments more effectively than Wavelet thresholding.

For artifact reduction on true artifact windows (*n* = 393), we assessed both methods. Wavelet thresholding achieved 18.8% amplitude reduction and 97.9% high-frequency (30–40 Hz) reduction, whereas EMD achieved 17.6% amplitude reduction and 99.8% high-frequency reduction (Table 2). The 1.9% difference in high-frequency reduction (EMD− Wavelet thresholding) reflects EMD’s enhanced ability to remove muscle artifacts in the 30–40 Hz band. EMD also achieved 99.1% beta-band (20–30 Hz) reduction compared to 87.0% for Wavelet thresholding. The 12.1% difference indicates stronger attenuation of muscle contamination across multiple frequency bands for EMD.

The difference in high-frequency reduction (99.8% for EMD versus 97.9% for Wavelet thresholding) suggests that removal of high-frequency intrinsic mode functions supports more complete artifact attenuation in the muscle artifact frequency range. Figures 4 and 5 show representative removal results for both methods.

**Figure 4:**
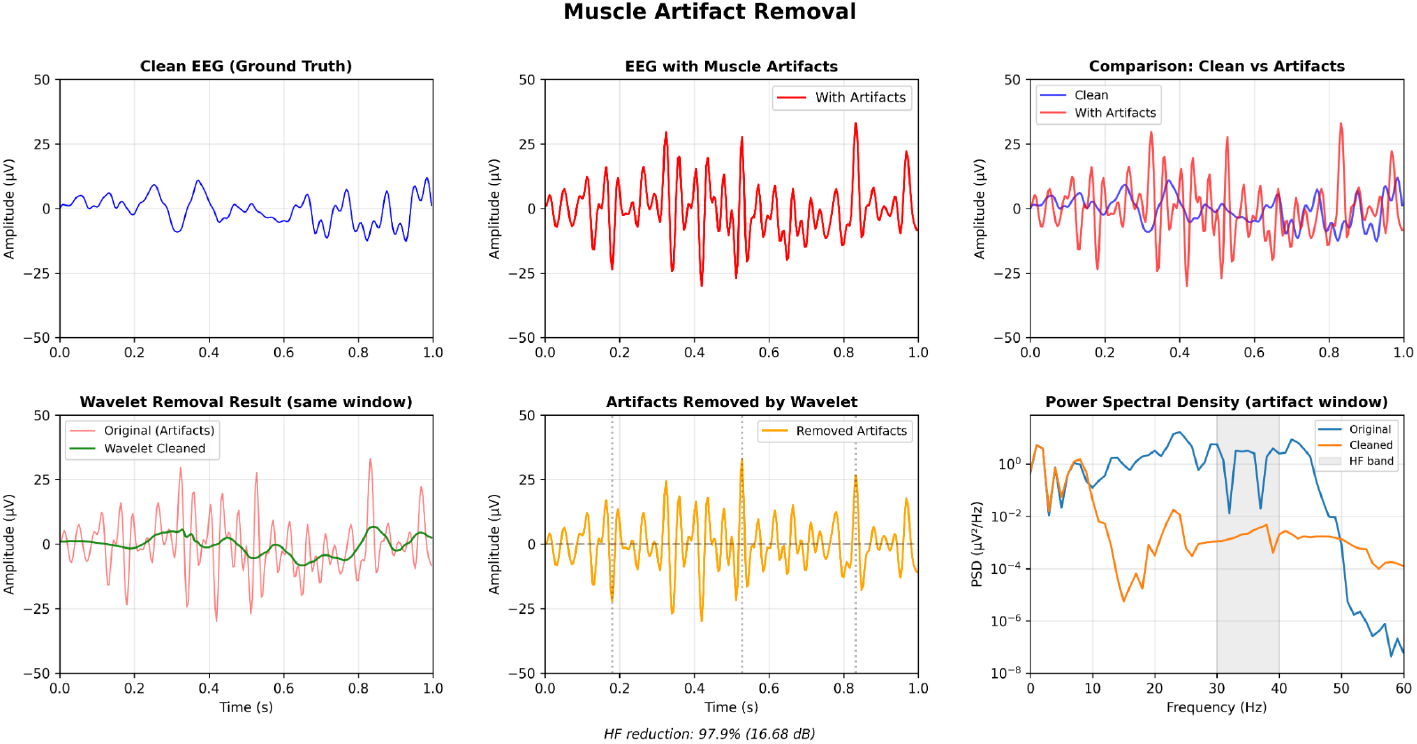
Wavelet muscle artifact removal: (top row) clean EEG reference window (separate artifact-free example), artifact-contaminated EEG, and comparison overlay; (bottom row) wavelet removal result (original vs cleaned) from the same detected artifact window, removed artifactual components, and power spectral density comparison showing HF band (30–40 Hz) reduction.

**Figure 5:**
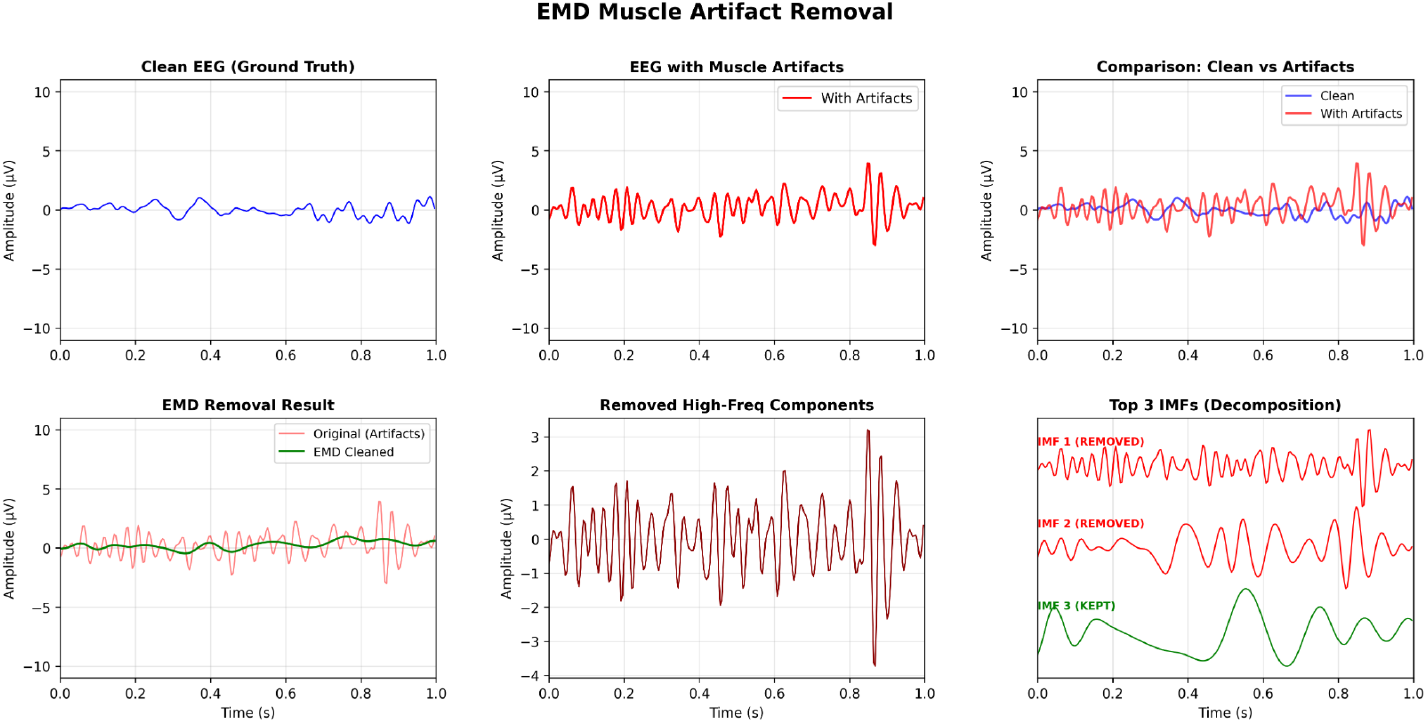
EMD muscle artifact removal: (top row) clean EEG reference window (separate artifact-free example), artifact-contaminated EEG, and comparison overlay; (bottom row) EMD removal result (original vs cleaned) from the same detected artifact window, removed high-frequency components, and top 3 IMFs showing which components were removed (IMFs 1–2) and kept (IMF 3).

#### Non-Physiological Artifact Removal

To evaluate non-physiological artifact removal performance, we tested spherical spline interpolation and ASR on the held-out test set (21 patients, 30 recordings). The CNN detector identified 169 artifact windows from 941 total windows and achieved an F1-score of 0.751 (precision 0.899, recall 0.644) (Table 3). The precision of 0.899 indicates that most detected windows contain true artifacts, and the recall of 0.644 indicates that the detector captures most artifact-contaminated segments. The F1-score of 0.751 indicates a balanced trade-off between precision and recall for non-physiological artifact detection.

**Table 3:** Non-physiological artifact removal summary on held-out test windows.

| Metric | Spline | ASR | $\Delta$<br>(ASR–Spline) |
| --- | --- | --- | --- |
| | $n = 941$ (236 artifact, 705 clean) | $n = 941$ (236 artifact, 705 clean) | |
|  | *Detected artifacts: 169* | *Detected artifacts: 169* |  |
|  | *Interpolated: 109* | *Processed: 169* |  |
|  | *Rejected: 60* | *Rejected: 0* |  |
| **Detection performance** |  |  |  |
| Precision | 0.899 | 0.899 | 0.000 |
| Recall | 0.644 | 0.644 | 0.000 |
| F1-score | 0.751 | 0.751 | 0.000 |
| **Clean preservation (clean windows, $n = 705$)** |  |  |  |
| PSNR (dB) | 68.44 | **69.64** | +1.20 |
| RMSE (μV) | 0.5045 | **0.4393** | -0.0652 |
| Correlation | 0.9873 | **0.9904** | +0.0031 |
| **Artifact reduction (processed windows)** |  |  |  |
| Amplitude reduction (%) | 26.6 | **37.1** | +10.5 |
| Peak reduction (%) | **49.6** | 47.2 | -2.4 |
| **Data retention** |  |  |  |
| Retention rate (%) | 93.6 | **100.0** | +6.4 |

**Table description:** Total windows: 941 (236 ground-truth artifact, 705 clean). Window length = 1 s (250 samples at 250 Hz). Removal is applied only to detected windows (no global correction). Spline: spherical spline interpolation reconstructing bad channels from spatial neighbors (Z-score thresholds: 2.0 SD variance, 1.8 SD amplitude; rejection threshold: 50% bad channels). ASR: artifact subspace reconstruction attenuating high-variance components (burst threshold: 2.5 SD). $\Delta$ reports ASR–Spline. For RMSE.

For clean-window preservation, we evaluated both methods. Spherical spline interpolation showed correlation of 0.987, RMSE of 0.504 *µ*V, and PSNR of 68.44 dB, whereas ASR showed correlation of 0.990, RMSE of 0.439 *µ*V, and PSNR of 69.64 dB (Table 3). Both methods showed correlation values above 0.987. These results indicate that the cleaned signals match the original clean segments with minimal distortion. The differences in PSNR (1.20 dB) and RMSE (0.065 *µ*V) indicate that ASR preserved clean segments more effectively than spherical spline interpolation.

For artifact reduction on processed windows, we assessed both methods. Spherical spline interpolation achieved 26.6% amplitude reduction and 49.6% peak reduction with 93.6% data retention, whereas ASR achieved 37.1% amplitude reduction and 47.2% peak reduction with 100.0% data retention (Table 3). The 37.1% reduction was computed on detector-flagged non-physiological artifact windows and reflects attenuation of high-amplitude non-physiological transients. Artifact-free windows showed minimal alteration under ASR (correlation 0.990, PSNR 69.64 dB). The 10.5% difference in amplitude reduction (ASR −spherical spline interpolation) reflects ASR’s enhanced ability to attenuate high-variance components across detected windows. ASR processed all 169 detected windows without rejection, whereas spherical spline interpolation rejected 60 windows (35.5%) that exceeded the contamination threshold. The difference in data retention (100.0% for ASR and 93.6% for spherical spline interpolation) indicates that ASR preserved a larger fraction of the recording.

The difference in data retention suggests that ASR can process all detected windows without rejection through attenuation of high-variance components. Figures 6 and 7 show representative removal results for both methods.

**Figure 6:**
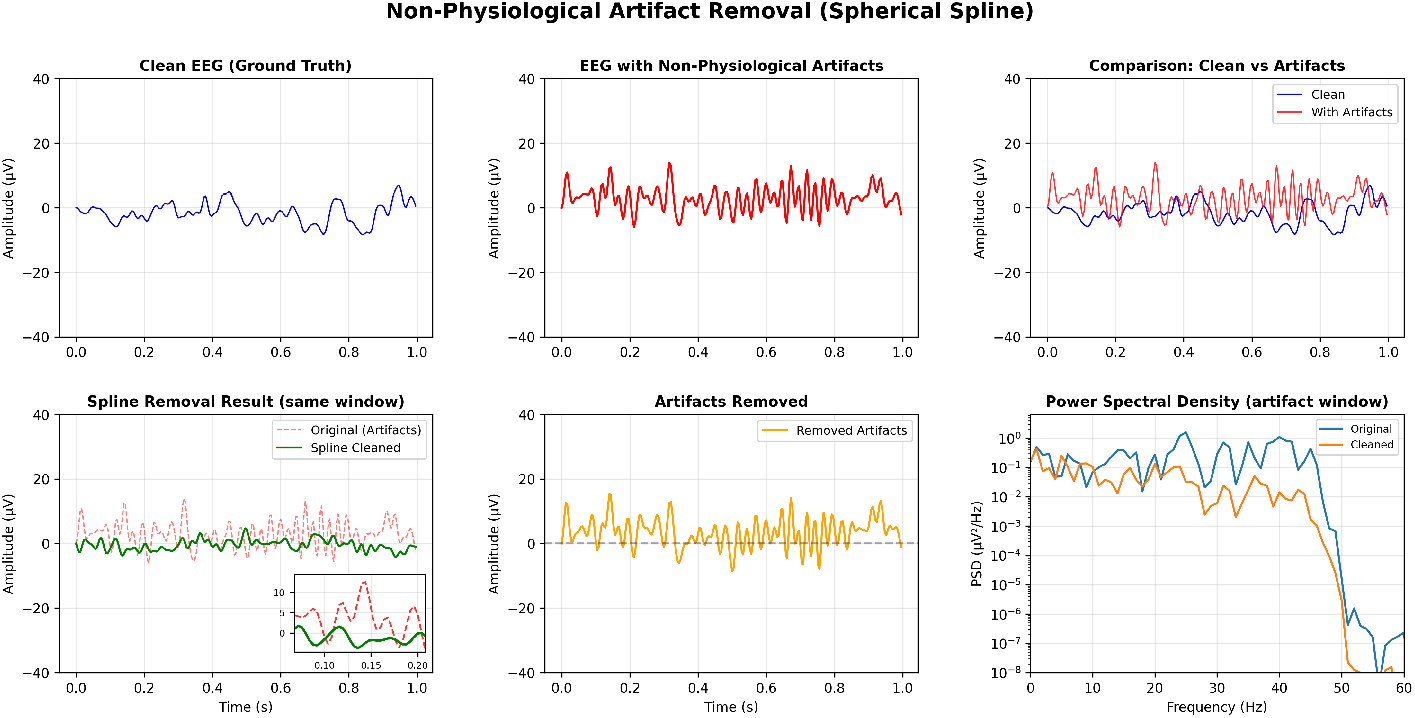
Spherical spline interpolation non-physiological artifact removal: (top row) clean EEG reference window (separate artifact-free example), artifact-contaminated EEG, and comparison overlay; (bottom row) spline removal result (original vs cleaned) from the same detected artifact window with zoom inset, removed artifactual components, and power spectral density comparison showing reduction across frequency bands.

**Figure 7:**
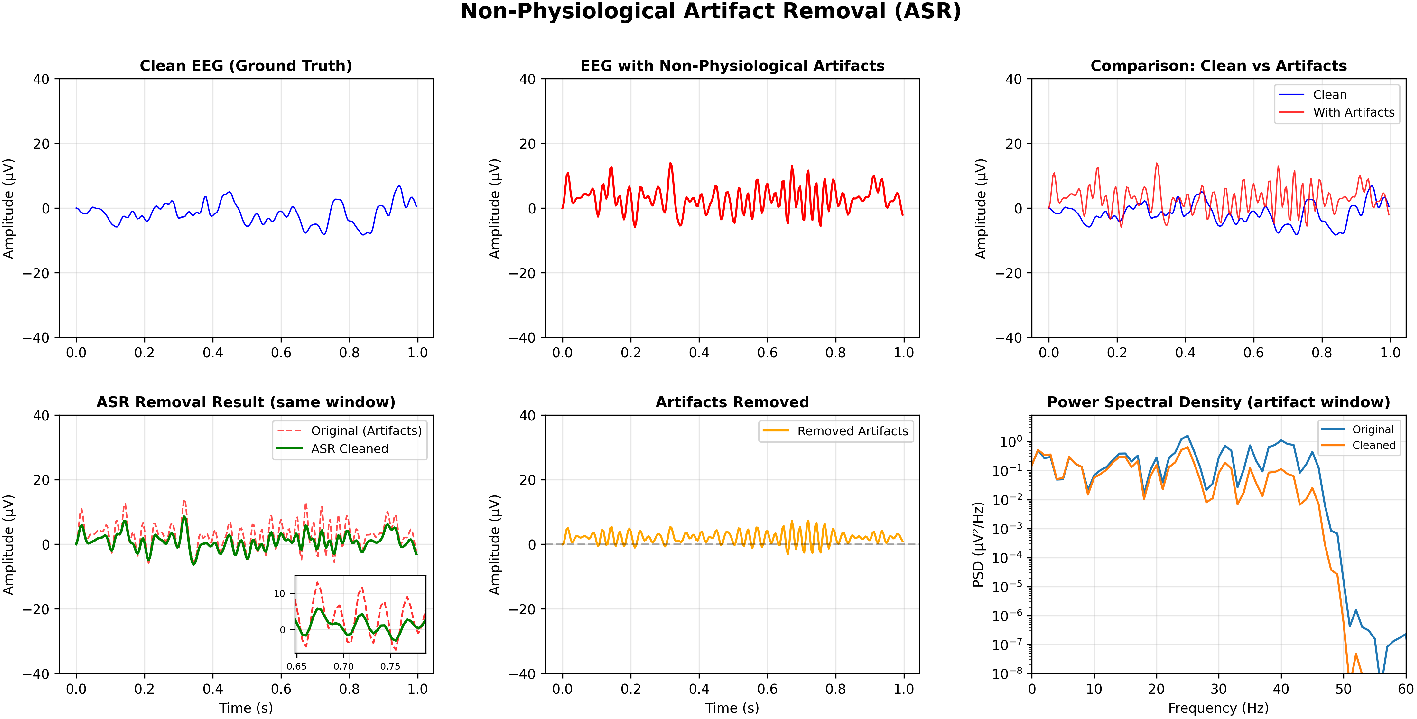
Artifact subspace reconstruction (ASR) non-physiological artifact removal: (top row) clean EEG reference window (separate artifact-free example), artifact-contaminated EEG, and comparison overlay; (bottom row) ASR removal result (original vs cleaned) from the same detected artifact window with zoom inset showing detailed artifact attenuation, removed artifactual components, and power spectral density comparison showing reduction across frequency bands.

### 3.2 Comparative Analysis

To evaluate the benefit of detection-guided selective removal, we compared selective and global removal strategies across all artifact types and methods (Table 4). For eye movement artifact removal, selective processing consistently improved clean-segment preservation relative to global removal. ICA showed a correlation of 0.9997 (selective) versus 0.8622 (global). These values indicate minimal distortion under selective removal and substantial alteration of clean windows under global removal. RMSE increased from 0.0827 *µ*V (selective) to 1.6117 *µ*V (global), and PSNR decreased from 84.15 dB (selective) to 58.35 dB (global). CCA showed a larger degradation under global removal. Correlation decreased from 0.9992 (selective) to 0.3899 (global). These results show severe distortion when CCA is applied indiscriminately to clean windows.

**Table 4:**
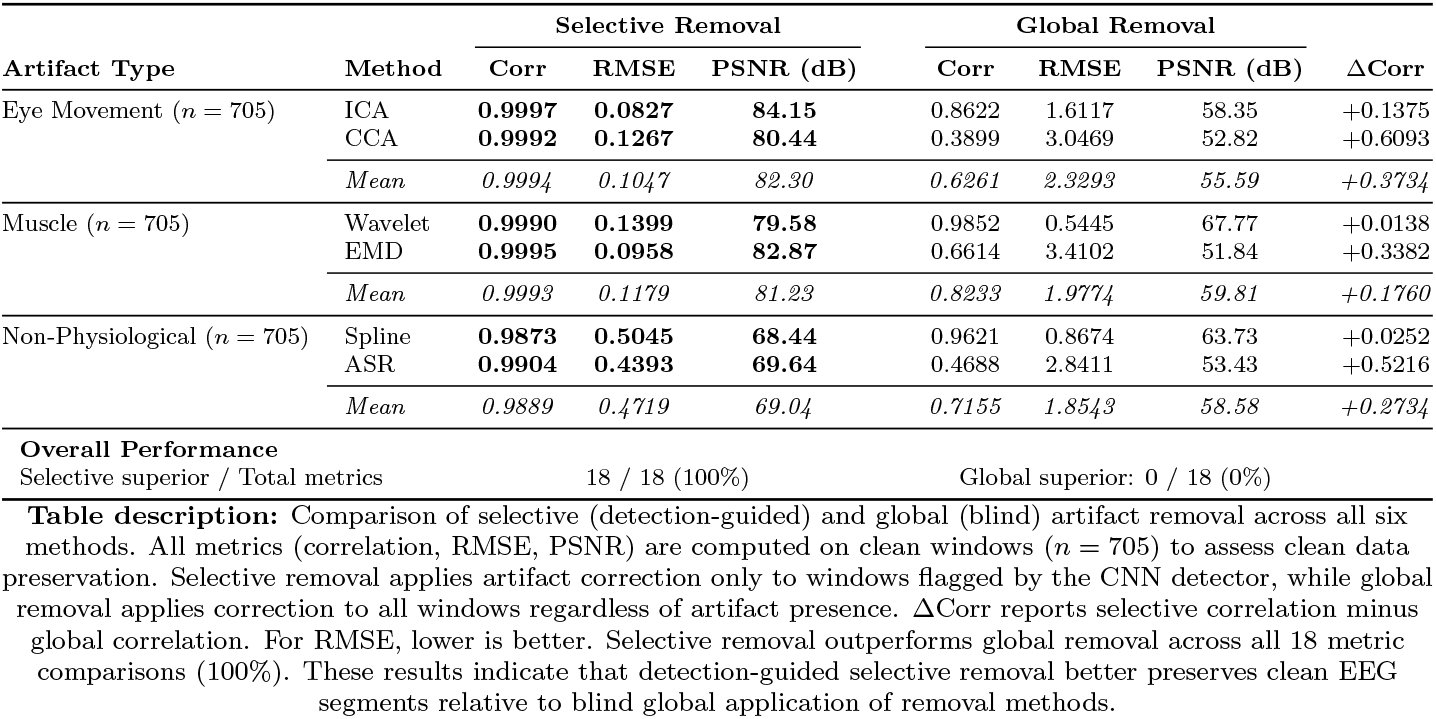
Comparative analysis of selective (detection-guided) and global (blind) artifact removal across all methods.

For muscle artifact removal, selective removal showed consistent advantages across both methods. Wavelet thresholding showed correlation of 0.9990 (selective) and 0.9852 (global). RMSE increased from 0.1399 *µ*V (selective) to 0.5445 *µ*V (global). EMD showed a larger effect.

Correlation decreased from 0.9995 (selective) to 0.6614 (global). These values show substantial distortion under global processing. PSNR differences showed the same pattern across both methods. For Wavelet thresholding, PSNR was 79.58 dB (selective) and 67.77 dB (global). For EMD, PSNR was 82.87 dB (selective) and 51.84 dB (global). These results show that selective removal maintains signal quality more effectively than global removal.

For non-physiological artifact removal, selective removal also improved clean-segment preservation, and the magnitude of improvement varied by method. Spherical spline interpolation showed correlation of 0.9873 (selective) and 0.9621 (global). RMSE increased from 0.5045 *µ*V (selective) to 0.8674 *µ*V (global). ASR showed severe degradation under global removal.

Correlation decreased from 0.9904 (selective) to 0.4688 (global). PSNR differences supported the same conclusion. For spherical spline interpolation, PSNR was 68.44 dB (selective) and 63.73 dB (global). For ASR, PSNR was 69.64 dB (selective) and 53.43 dB (global).

Across all six removal methods and 18 metric comparisons, selective removal outperformed global removal in 100% of cases. These consistent improvements in correlation, RMSE, and PSNR indicate that detection-guided selective removal better preserves clean EEG segments than blind global application of artifact removal methods.

## 4. Discussion

In this study, we evaluated a detection-guided artifact removal framework for clinical EEG. The framework applies removal methods selectively to detected artifact windows while preserving clean segments unchanged. This selective approach addresses a critical limitation of existing pipelines that apply correction globally. Our evaluation included (a) implementation of artifact-specific removal methods guided by CNN detection outputs, (b) comparison of selective and global removal strategies to quantify the benefit of detection-guided processing, and (c) assessment of clean data preservation to ensure clinical safety. The framework achieved strong performance across all three artifact types and six removal methods. Selective removal outperformed global removal across all 18 metric comparisons.

### 4.1 Advantages of Detection-Guided Selective Removal

The key methodological choice in our framework is the selective application of removal methods based on detection outputs rather than global application across all windows. Most existing pipelines apply removal globally across entire recordings or uniformly across all artifact types[12, 13, 33]. This global approach risks over-aggressive processing of clean segments. Our comparative analysis showed that global removal introduced substantial distortion to clean windows. Correlation dropped to 0.3899 for CCA and 0.4688 for ASR. The selective approach avoids this distortion. It applies removal only to detected artifact windows and preserves clean segments unchanged.

The detection-guided approach addresses a limitation of existing methods that assume homogeneous artifact distributions across channels, subjects, and tasks[38, 39, 40]. The CNN detectors adapt to the specific characteristics of each recording through training on diverse clinical data. The detection-guided removal applies methods only where artifacts are present. This approach reduces the risk of signal distortion in clean segments and maintains effective artifact suppression in contaminated segments. The modular design allows integration of different removal methods for each artifact type. This design supports tailored approaches based on artifact characteristics.

The framework processes artifacts automatically based on CNN detection outputs. This automation eliminates the need for manual artifact identification and method selection. Selective application ensures that removal methods are used only where needed. Targeted processing reduces redundant operations and maintains signal quality in clean segments. The automated workflow addresses the workload burden identified in existing studies[12, 13, 14]. Manual artifact review requires extensive time and expertise in those settings.

### 4.2 Effect of Selective Removal on Clean Data Preservation

The comparative analysis demonstrated that detection-guided selective removal outperformed global removal across all six methods and 18 metric comparisons. Selective removal maintained correlation above 0.987 for all methods, whereas global removal showed correlation as low as 0.3899 for CCA and 0.4688 for ASR. Statistical analysis confirmed that selective removal produced lower RMSE than global removal for all six methods (*p*-values ranging from 2.54 *×* 10^−105^ to 4.42 *×* 10^−116^). The 100% success rate across all methods and metrics demonstrates that detection-guided selective removal is superior to blind global application of removal methods.

For eye movement artifacts, selective ICA maintained correlation of 0.9997 and selective CCA maintained 0.9992, whereas global removal degraded these values to 0.8622 (ICA) and 0.3899 (CCA). The correlation drop from 0.9992 to 0.3899 for CCA reflects the cost of processing clean windows that do not require artifact removal. For muscle artifacts, selective Wavelet thresholding maintained correlation of 0.9990 and selective EMD maintained 0.9995, whereas global removal degraded these values to 0.9852 (Wavelet thresholding) and 0.6614 (EMD). The correlation dropped from 0.9995 to 0.6614 for EMD, indicating that global removal causes substantial distortion when applied to all windows.

For non-physiological artifacts, selective spherical spline interpolation maintained correlation of 0.9873 and selective ASR maintained 0.9904, whereas global removal degraded these values to 0.9621 (spherical spline interpolation) and 0.4688 (ASR). The correlation dropped from 0.9904 to 0.4688 for ASR, indicating that global removal causes severe distortion when applied to all windows. PSNR differences show similar patterns. Selective removal maintained PSNR above 68 dB for all methods, while global removal showed PSNR as low as 51.84 dB for EMD and 53.43 dB for ASR.

### 4.3 Performance Across Artifact Types and Removal Methods

The framework achieved strong performance across all three artifact types and six removal methods on the held-out test set. Both ICA and CCA achieved excellent clean segment preservation (correlation *>* 0.999, PSNR *>* 80 dB). The methods differed substantially in artifact reduction. CCA removed 74.6% of amplitude compared to 35.0% for ICA. The 39.6% difference reflects CCA’s ability to extract correlated components between frontal and non-frontal channels. The high correlation values indicate minimal distortion under selective application.

Wavelet thresholding and EMD showed similarly strong preservation (correlation *>* 0.999, PSNR *>* 79 dB). EMD demonstrated superior frequency-specific reduction. EMD achieved 99.8% high-frequency reduction compared to 97.9% for Wavelet thresholding, and EMD achieved 99.1% beta band reduction compared to 87.0% for Wavelet thresholding. The 12.1% difference in beta band reduction shows that EMD more effectively attenuates muscle contamination across multiple frequency bands. These preservation metrics confirm minimal distortion under selective application.

Spherical spline interpolation and ASR both maintained high preservation quality (correlation *>* 0.987, PSNR *>* 68 dB) but differed in their approach to ambiguous windows. ASR processed all 169 detected windows without rejection (100% retention), whereas spherical spline interpolation rejected 60 windows (35.5%) that exceeded contamination thresholds. ASR achieved 37.1% amplitude reduction compared to 26.6% for spherical spline interpolation. The 10.5% difference reflects ASR’s enhanced ability to attenuate high-variance components across all detected windows. The choice between methods depends on the clinical priority. Spherical spline interpolation provides conservative processing with rejection of ambiguous cases, whereas ASR provides complete data retention with aggressive artifact attenuation.

### 4.4 Clinical Feasibility and Safety

The framework has direct implications for clinical EEG monitoring in acute and perioperative care settings. Continuous EEG monitoring requires rapid artifact identification and removal to support real-time seizure detection and therapeutic decision-making[6, 7, 8]. The detection-guided approach enables automated artifact removal without manual review. This approach reduces reviewer workload and improves diagnostic precision[12, 13, 14]. Selective application of removal methods preserves artifact-free segments without modification. This preservation helps ensure that genuine pathology is not masked by false corrections[15, 9].

Preservation of artifact-free segments with correlation above 0.987 and PSNR above 68 dB across all methods indicates that the framework maintains clinical safety. The removal methods preserve waveform morphology and spectral content within acceptable limits for clinical interpretation. Statistical analysis confirmed that selective removal better preserves clean data than global removal. These results support the clinical feasibility of the detection-guided approach. The framework can be integrated into existing EEG monitoring systems to provide automated artifact removal without compromising signal quality. Artifact attenuation occurred in detector-flagged windows, and preservation metrics on artifact-free windows indicated minimal distortion of clinically relevant EEG features.

Data retention rates of 93.6% for spherical spline interpolation and 100% for ASR indicate that the framework preserves most of the recording for clinical analysis. Rejection of severely contaminated windows by spherical spline interpolation ensures that only high-quality data are retained, whereas ASR processes all detected windows without rejection. The preservation metrics are consistent with clinical safety requirements for maintaining waveform morphology and spectral content in artifact-free segments. This safety is important for clinical adoption, where false corrections could mask genuine pathology[6, 7, 14].

### 4.5 Limitations and Future Directions

Several limitations should be considered when interpreting our results. The evaluation was conducted on a single dataset (TUH Artifact Corpus) with specific recording conditions and patient populations. The framework may require validation on other datasets with different equipment, electrode configurations, and clinical settings to ensure generalizability[12, 13, 33]. The detection models were trained on data from 2002 to 2016, and performance may vary with newer recording systems or different artifact characteristics.

The framework relies on the accuracy of the CNN detectors, and detection errors can propagate into removal performance. False positive detections may lead to unwarranted processing of clean segments, whereas false negative detections may leave artifacts unremoved. Detection performance showed F1-scores ranging from 0.751 to 0.895 across artifact types. This range indicates room for improvement. Future work should prioritize higher detection accuracy and lower false positive and false negative rates.

The removal methods were evaluated independently for each artifact type, but real-world recordings often contain multiple artifact types simultaneously. The framework processes artifacts sequentially by type, which may not account for interactions between different artifact sources. The evaluation did not assess performance on recordings with overlapping artifacts or complex artifact combinations. Future work should evaluate the framework on recordings with multiple simultaneous artifacts to assess robustness in complex scenarios.

Several directions for future work could improve the framework. Detection performance could be improved through ensemble methods, transfer learning from other datasets, or adaptive thresholding based on recording characteristics[34, 35, 36]. The removal methods could be optimized for specific clinical applications, such as seizure detection or sleep staging, where different artifact characteristics may require tailored approaches[6, 7]. The framework could be extended to handle additional artifact types, such as cardiac artifacts, electrode drift, or environmental noise.

Real-time implementation would enable integration into continuous monitoring systems. This integration requires improved computational efficiency and reduced latency[8]. Validation on diverse datasets from different institutions, recording systems, and patient populations would strengthen the generalizability of the framework. Prospective clinical studies could assess the impact of automated artifact removal on diagnostic accuracy, reviewer workload, and patient outcomes[15, 9]. The open-source release of the framework will facilitate community validation and adoption in clinical and research settings.

## 5. Conclusion

We developed and validated a detection-guided artifact removal framework that selectively processes contaminated EEG segments while preserving clean data unchanged. Across all six removal methods and three artifact types, selective processing outperformed global removal in all 18 preservation metrics tested (*p <* 10^−105^). Clean segment correlation remained above 0.987, whereas global methods produced values as low as 0.39. The framework achieved effective artifact suppression and preserved clinical signal quality. ICA and CCA removed up to 74.6% of eye movement artifact amplitude. Wavelet thresholding and EMD removed up to 99.8% of high-frequency muscle contamination. Spherical spline interpolation and ASR reduced non-physiological artifact amplitude by up to 37.1%. Data retention rates ranged from 93.6–100%.

The modular, automated design eliminates manual artifact review and enables integration of tailored removal methods for specific artifact characteristics.

## Acknowledgments

The authors thank the Temple University Hospital for providing access to the TUH EEG Corpus and the clinical neurophysiology team for valuable feedback on algorithm validation.

## Author contributions

E.N.: Conceptualization, Methodology, Software, Validation, Formal analysis, Investigation, Data curation, Writing - original draft, Writing - review & editing, Visualization. P.D.T.: Supervision, Validation, Writing - review & editing, Resources. S.V.: Supervision, Writing - review & editing. Z.G.: Supervision, Methodology, Formal analysis, Writing - review & editing, Project administration.

## Data availability

All code, trained models, and evaluation scripts will be made publicly available at [GitHub repository] upon publication. The TUH EEG Artifact Corpus is available through the Temple University Hospital EEG Corpus at https://isip.piconepress.com/projects/nedc/html/tuh_eeg/. Access requires completion of a data use agreement form submitted to.

## References

[1] Bitar R, Khan U M and Rosenthal E S 2024 Critical Care 28 244

[2] Limotai C, Ingsathit A, Thadanipon K, McEvoy M, Attia J and Thakkinstian A 2019 Critical care medicine 47 e366–e373

[3] Holla S K, Krishnamurthy P V, Subramaniam T, Dhakar M B and Struck A F 2022 Neurologic clinics 40 907–925

[4] Kubota Y, Nakamoto H, Egawa S and Kawamata T 2018 Journal of Intensive Care 6 39

[5] Swisher C B, Shah D, Sinha S R and Husain A M 2015 Journal of Clinical Neurophysiology 32 147–151

[6] Sharma S, Nunes M and Alkhachroum A 2022 Frontiers in Neurology 13 951286

[7] Rossetti A O, Claassen J and Gaspard N 2024 Intensive care medicine 50 1–16

[8] Fenter H, Rossetti A O and Beuchat I 2024 European neurology 87 17–25

[9] Claassen J, Mayer S A, Kowalski R G, Emerson R G and Hirsch L J 2004 Neurology 62 1743–1748

[10] DeLorenzo R, Hauser W, Towne A, Boggs J, Pellock J, Penberthy L, Garnett L, Fortner C and Ko D 1996 Neurology 46 1029–1035

[11] Young B G, Jordan K G and Doig G S 1996 Neurology 47 83–89

[12] Islam M K, Rastegarnia A and Yang Z 2016 Neurophysiologie Clinique 46 287–305

[13] Uriguen J A and Garcia-Zapirain B 2015 Journal of Neural Engineering 12 031001

[14] Amin U, Nascimento F A, Karakis I, Schomer D and Benbadis S R 2023 Epileptic disorders 25 591–648

[15] Alkhachroum A, Appavu B, Egawa S, Foreman B, Gaspard N, Gilmore E J, Hirsch L J, Kurtz P, Lambrecq V, Kromm J et al. 2022 Intensive care medicine 48 1443–1462

[16] Goncharova I I, McFarland D J, Vaughan T M and Wolpaw J R 2003 Clinical Neurophysiology 114 1580–1593

[17] Whitham E M, Pope K J, Fitzgibbon S P, Lewis T, Clark C R, Loveless S, Broberg M, Wallace A, DeLosAngeles D, Lillie P et al. 2007 Clinical Neurophysiology 118 1877–1888

[18] Muthukumaraswamy S D 2013 Frontiers in human neuroscience 7 138

[19] Nolan H, Whelan R and Reilly R B 2010 Journal of Neuroscience Methods 192 152–162

[20] Delorme A and Makeig S 2004 Journal of Neuroscience Methods 134 9–21

[21] Chen X, Liu A, Chiang J, Wang Z J, McKeown M J and Ward R K 2015 IEEE Sensors Journal 16 1986–1997

[22] Shahbakhti M, Khalili V and Kamaee G 2012 Removal of blink from EEG by empirical mode decomposition (EMD) The 5th 2012 Biomedical Engineering International Conference (IEEE) pp 1–5

[23] Kiamini M, Alirezaee S, Perseh B and Ahmadi M 2009 Elimination of ocular artifacts from eeg signals using the wavelet transform and empirical mode decomposition 2009 6th International Conference on Electrical Engineering/Electronics, Computer, Telecommunications and Information Technology vol 2 (IEEE) pp 1094–1097

[24] Dai Y, Duan F, Feng F, Sun Z, Zhang Y, Caiafa C F, Marti-Puig P and Solé-Casals J 2021 Entropy 23 1170

[25] Phadikar S, Sinha N and Ghosh R 2020 IEEE Journal of Biomedical and Health Informatics 25 475–484

[26] Saini M, Satija U and Upadhayay M D 2020 IEEE Signal Processing Letters 27 1260–1264

[27] Cataldo A, Criscuolo S, Benedetto E D, Masciullo A, Pesola M, Schiavoni R and Invitto S 2022 IEEE Sensors Journal 22 21257–21265

[28] Plechawska-Wojcik M, Kaczorowska M and Zapala D 2018 The artifact subspace reconstruction (asr) for eeg signal correction. a comparative study International conference on information systems architecture and technology (Springer) pp 125–135

[29] Dong L, Zhao L, Zhang Y, Yu X, Li F, Li J, Lai Y, Liu T and Yao D 2021 Brain Topography 34 403–414

[30] Peng H, Hu B, Shi Q, Ratcliffe M, Zhao Q, Qi Y and Gao G 2013 IEEE journal of biomedical and health informatics 17 600–607

[31] Abidi A, Nouira I, Assali I, Saafi M A and Bedoui M H 2022 The Computer Journal 65 1257–1271

[32] Mowla M R, Ng S C, Zilany M S and Paramesran R 2015 Biomedical Signal Processing and Control 22 111–118

[33] Molina-Molina M, Tardón LJ, Barbancho A M and Barbancho I 2024 Applied Sciences 14 971

[34] Kalita B, Deb N and Das D 2024 Scientific Reports 14 24234

[35] Jiang R, Tong S, Wu J, Hu H, Zhang R, Wang H, Zhao Y, Zhu W, Li S and Zhang X 2025 Scientific Reports 15 19419

[36] Xiong W, Ma L and Li H 2024 Frontiers in Neuroscience 17 1258024

[37] Sawangjai P, Trakulruangroj M, Boonnag C, Piriyajitakonkij M, Tripathy R K, Sudhawiyangkul T and Wilaiprasitporn T 2021 IEEE Journal of Biomedical and Health Informatics 26 4913–4924

[38] Roy Y, Banville H, Albuquerque I, Gramfort A, Falk T H and Faubert J 2019 Journal of Neural Engineering 16 051001

[39] Craik A, He Y and Contreras-Vidal J L 2019 Journal of Neural Engineering 16 031001

[40] Mvvs P R T 2024 Digital Signal Processing

[41] Mathe M, Mididoddi P and Krishna B T 2022 Proceedings of Engineering and Technology Innovation 20 35–56

[42] Nyanney E, Thirumala P D, Visweswaran S and Geng Z 2025 medRxiv 2025–10

[43] Krishnaveni V, Jayaraman S, Anitha L and Ramadoss K 2006 Journal of Neural Engineering 3 338–346

[44] Winkler I, Haufe S and Tangermann M 2011 Behavioral and Brain Functions 7 30

[45] Clercq W D, Vergult A, Vanrumste B, Paesschen W V and Huffel S V 2006 IEEE Transactions on Biomedical Engineering 53 2583–2587

[46] Obeid I and Picone J 2016 Frontiers in Neuroscience 10 196

[47] López S 2020 The tuh eeg artifact corpus https://isip.piconepress.com/projects/tuh_eeg/

[48] Perrin F, Pernier J, Bertrand O and Echallier J F 1989 Electroencephalography and clinical neurophysiology 72 184–187

